# Gendered sport environments and their theoretical contributions to women’s ACL injury risk: a scoping review

**DOI:** 10.1101/2024.10.09.24315187

**Authors:** Joanne L Parsons, Olayinka Akinrolie, Hal Loewen, Stephanie E Coen

## Abstract

**Objective:** To summarize the current literature that describes the gendered environments of women’s sports which are higher risk for ACL injury, and determine whether the existing literature relates gendered aspects of the sport environment to injury.

**Design:** Scoping review.

**Data sources:** Electronic search of Medline, Embase, CINAHL, SPORTDiscus, SCOPUS, and Women’s Studies International databases from inception to March 2024.

**Eligibility criteria:** Studies were included if at least 50% of the study participants were adult women participating in organized sports with higher risk for ACL injury.

**Results:** Of the initially identified 17,148 studies, 854 underwent full text review, and 73 were included in this scoping review. In 19 studies, reference to injury was restricted to one or two direct quotes from an athlete and the other 54 studies had no mention of injury. We identified three repeating patterns describing the gendered sport environments that women athletes encounter. Fifty-five studies described embedded stereotypes that devalue women and women’s sport. Forty-five studies described ways the sport environment reproduces restrictive gender norms for women. Forty-six studies reported that gendered inequities including gendered wage inequality and provision of subpar training facilities were structurally embedded in women’s sport environments.

**Conclusion:** Existing literature describes a range of gendered inequities that exist for women in their sport environments; however, there has been no concerted effort to date to link those gendered environmental factors to ACL injury. Such research is needed if we are serious about eliminating the ACL injury rate disparity between women and men.

**What is already known:**

- Historically, gendered disparities in anterior cruciate ligament injury risk have focused on biological explanations
- Societal gendered norms and expectations of women can greatly affect their experiences and opportunities in sport environments

**What are the new findings:**

- Existing literature describes a range of gendered inequities that exist for women in their sport environments
- Only a small percentage (∼25%) of studies describing the gendered aspects of women’s sport environments mention any relation to injury
- There has been no concerted effort to link gendered aspects of sport environments to women’s injury risk and experiences

## INTRODUCTION

Sport participation offers numerous benefits, including improved physical and mental health, social connection, and a sense of community.^1^ However, because sport opportunities were historically created for men, by men, gendered inequities are arguably an inevitable part of the sport system. This can adversely affect participation rates and the experiences that girls and women have in sport.^2^ The negative consequences may even extend to physical injury risk, as girls and women experience anterior cruciate ligament (ACL) injuries at significantly higher rates compared to boys and men.^3,4^ The consequences of these injuries can be severe. Sustaining an ACL injury can lead to early development of osteoarthritis and the need for joint replacement^5^ and has lifelong implications for health and physical activity engagement for the athlete.^6,7^ In high performance environments where training and competition are often structured around a rigid 4-year Olympic cycle, loss of player availability can compromise the sport organization’s performance targets, and athletes can be at risk of losing critical funding.

Attempts to explain and remedy ACL injury rate disparities between women and men have met with limited success over the last 30 years.^8,9^ One significant factor in the lack of progress may be the overwhelming focus on biological explanations for injury risk (e.g., anatomy, menstrual cycle).^10–13^ Possible wider societal influences have rarely been considered, and so the majority of risk factors for injury are considered intrinsic to the athlete and their individual responsibility to address. However, wider societal gendered norms and expectations of girls and women have been shown to greatly affect their experiences, opportunities, and participation in sport environments.^14,15^ One clear example is the weight room environment – while resistance training is a key component for effective sports injury prevention and rehabilitation, many girls and women feel uncomfortable and even unwelcome in those spaces.^16–19^ Work in the public health arena suggests that injury prevention efforts are most effective when broader society and cultural norms are targeted, rather than placing the burden on individual people to change their behaviour.^20,21^ To reconceptualize opportunities to intervene in sports injury from this widened lens, we previously developed a gendered environmental approach to illustrate how gender can operate as an extrinsic determinant of risk from the pre-sport, training, and competition environments through to injury and the treatment environment (Figure 1).^22^ To date, the field of sports medicine has not approached sports injury from this broader perspective, and we lack critical information regarding the gendered features of the sport environment that may be contributing to increased injury risk for women.

**Figure 1.**
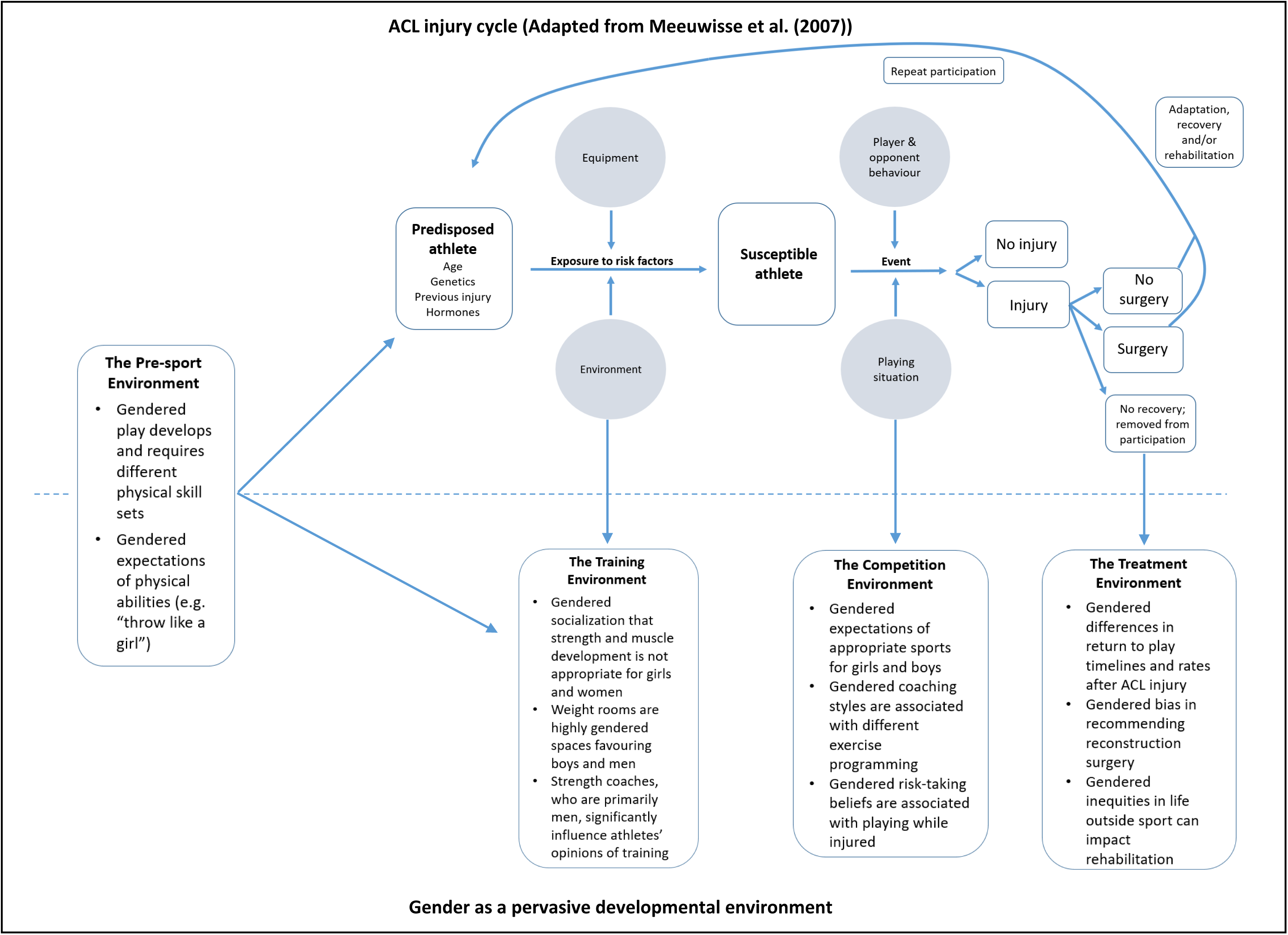
Gender as a pervasive developmental environment in the injury cycle. Reproduced from Anterior cruciate ligament injury: towards a gendered environmental approach, Parsons JL, Coen SE, Bekker S, 55:984-990;2021, with permission from BMJ Publishing Group Ltd.

This scoping review primarily sought to answer the question: What literature currently exists that describes the gendered aspects of women’s sports environments which are higher risk for ACL injury? We also sought to identify whether this existing literature relates gendered aspects of the sports environment specifically to injury; determine whether the described gendered sports environments align with the 4 environments depicted in our gendered environmental approach to sports injury; and summarize the gendered environments that exist for women in sport.

## METHODS

### Equity, diversity, and inclusion statement

Our research team is gender balanced, spans career stages from a recent PhD graduate to mid and late-career academics, and draws together expertise across social sciences, clinical health sciences, and library sciences. Our study set out to identify inequities for women in sport, and in the data collection phase we purposefully included a column in our extraction table to capture mention of any intersectionality (e.g., race/ethnicity, religion, sexuality) in the included papers.

### Study design

We undertook a scoping review, as this type of review is especially useful in emerging research areas such as this in which a variety of methods are employed in the publications of interest.^23^ As we were designing the study, a preliminary search indicated that there were no similar existing reviews examining the relationship between the gendered aspects of women’s sport environments and injury. We registered the scoping review on Open Science Framework (https://osf.io/9b7kh). The theoretical framework guiding the project is outlined below, followed by a description of the stages for our scoping review which closely follow previous authors’ recommendations,^23–26^ and reflect the Preferred Reporting Items for Systematic reviews and Meta-Analyses extension for Scoping Reviews (PRISMA-ScR) guidelines (Supplementary File 1).^27^

### Theoretical framework

The historical approach to the study and management of sports injury decontextualizes sporting bodies from the world around us, instead focusing heavily on biological explanations. To reconsider sports injury from a wider lens, we developed a gendered environmental approach that views sports injury as a bio-*social* phenomenon where sporting bodies are situated in social environments that may have ‘gendering’ effects, to the detriment of women and girls.^22^ We argue that the dominant sports injury paradigms used to date not only foreclose other possible explanations for gendered inequities in injury risk and outcomes, but can also propagate sexist beliefs that women’s bodies are “unfit” for sport. Using the example of ACL injury, our model (Figure 1) illustrates how adding gender as a pervasive developmental environment^28^ requires us to consider the many plausible ways in which wider societal forces may influence sports injury risk and outcomes, and forces us to think beyond defaulting to biological differences. Our approach^22^ proposes that sport consists of four potentially gendered environments – 1) Pre-sport, 2) Training, 3) Competition, and 4) Treatment. Bringing this perspective to the sports medicine realm means we require the physical sporting body to be viewed as situated in social and material environments that, overtly or not, shape injury risk, prevention, and rehabilitation in highly gendered ways. We used our gendered environmental approach to sports injury to guide the collating, summarizing, synthesis, and reporting stages of our scoping review.

### Eligibility criteria

Inclusion and exclusion criteria were guided by the “Participants, Concept, Context” principles of scoping reviews^26^ and finalized through multiple research team discussions as the screening process unfolded (Table 1). Quantitative, qualitative, and mixed method designs were considered. The first author determined those sports included in the Paris 2024^29^ and Milan 2026^30^ Olympic Games that are higher risk for ACL and therefore suitable for inclusion.^4,31–44^ The list of included Olympic program sports can be found in Appendix A.

**Table 1.**
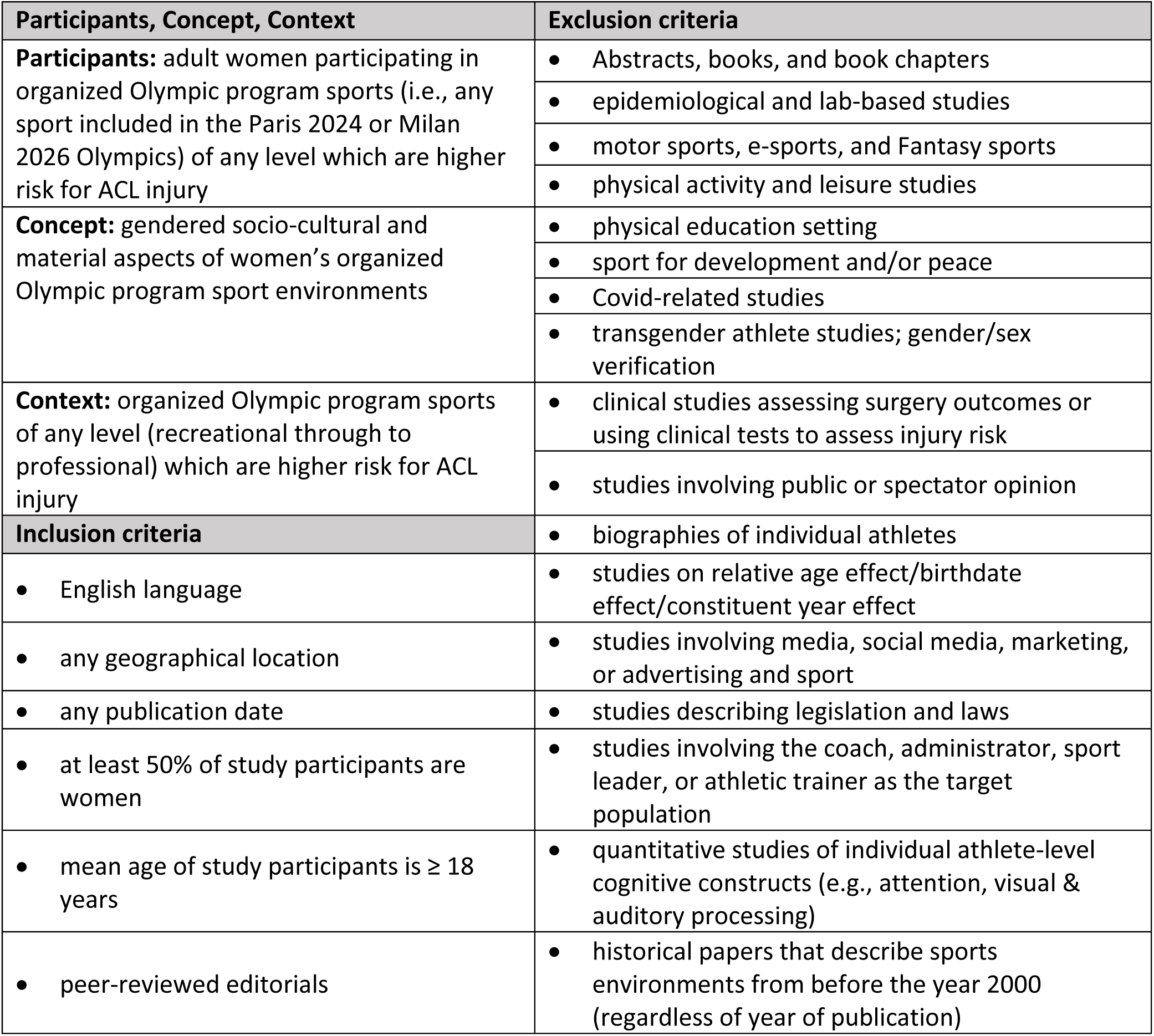
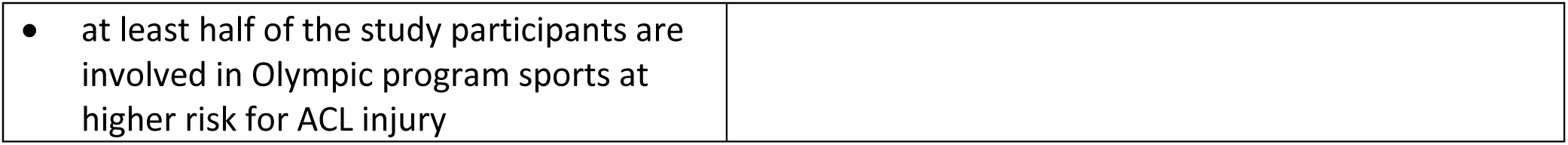
Inclusion and exclusion criteria.

| **Table 1. Inclusion and exclusion criteria** |  |
| --- | --- |
| **Participants, Concept, Context** | **Exclusion criteria** |
| **Participants:** adult women participating in organized Olympic program sports (i.e., any sport included in the Paris 2024 or Milan 2026 Olympics) of any level which are higher risk for ACL injury | • Abstracts, books, and book chapters<br>• epidemiological and lab-based studies<br>• motor sports, e-sports, and Fantasy sports<br>• physical activity and leisure studies |
| **Concept:** gendered socio-cultural and material aspects of women’s organized Olympic program sport environments | • physical education setting<br>• sport for development and/or peace<br>• Covid-related studies<br>• transgender athlete studies; gender/sex verification |
| **Context:** organized Olympic program sports of any level (recreational through to professional) which are higher risk for ACL injury | • clinical studies assessing surgery outcomes or using clinical tests to assess injury risk<br>• studies involving public or spectator opinion |
| **Inclusion criteria** | • biographies of individual athletes |
| • English language | • studies on relative age effect/birthdate effect/constituent year effect |
| • any geographical location | • studies involving media, social media, marketing, or advertising and sport |
| • any publication date | • studies describing legislation and laws |
| • at least 50% of study participants are women | • studies involving the coach, administrator, sport leader, or athletic trainer as the target population |
| • mean age of study participants is $\geq 18$ years | • quantitative studies of individual athlete-level cognitive constructs (e.g., attention, visual & auditory processing) |
| • peer-reviewed editorials | • historical papers that describe sports environments from before the year 2000 (regardless of year of publication) |

|  |
| --- |
| • at least half of the study participants are involved in Olympic program sports at higher risk for ACL injury |

### Data sources

In consultation with the Health Sciences librarian (HL) and using key terms from our primary research question and sub-questions, we created a comprehensive initial search strategy to use across multiple electronic databases. The initial search strategy (Appendix B) was peer reviewed by an additional health sciences librarian not involved with the study to ensure subject headings, keywords, and the search syntax were correct. The search was run across the following databases - Medline, Embase, CINAHL, SPORTDiscus, SCOPUS, and Women’s Studies International. The initial search was completed in December 2022 and identified approximately 10,000 potential studies. An updated search was completed in March 2024 to capture studies published after the initial search. Results were uploaded to Covidence (Veritas Health Innovation, Melbourne, Australia), a web-based software platform that facilitates the review process; duplicates were identified and removed. A total of 10,944 studies were identified from the database search that required screening (Figure 2).

**Figure 2.**
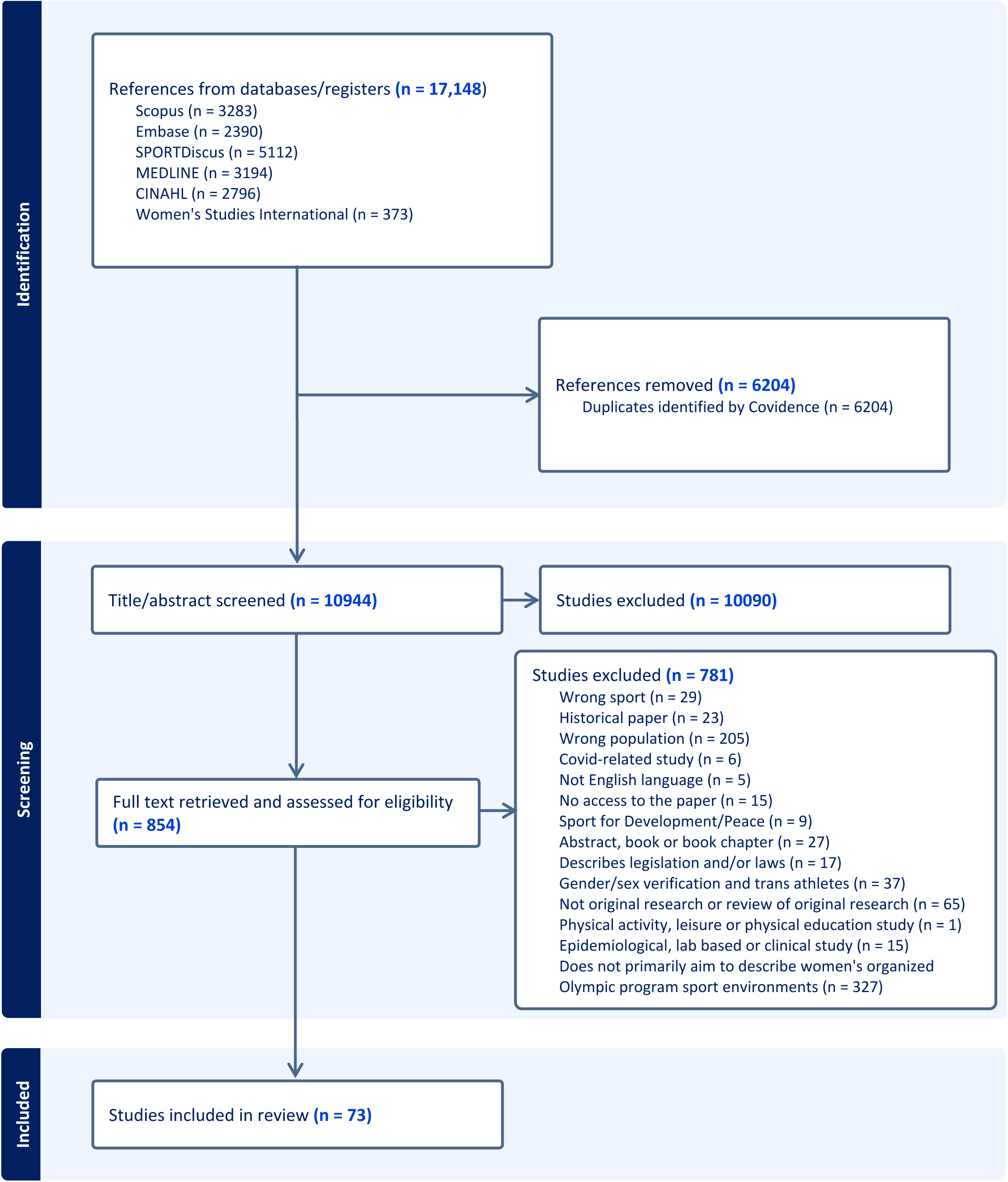
PRISMA flow diagram indicating study identification and selection.

### Study selection

To complete the initial title and abstract screening, every reference was screened by two research team members chosen at random by Covidence. We undertook three pilot sessions where the team screened approximately 50 titles each before meeting as a group to discuss the process and resolve conflicts. The research question was refined at this point to focus on Olympic sports that were high risk to ACL injury, and we refined our exclusion criteria. Once we were satisfied with the level of agreement among reviewers, we completed the initial screening, which included both title and abstract review. If the inclusion and exclusion criteria were met, or if it was unclear to the reviewers as to the relevance of an article just by reading the abstract, the study was moved to full text review.^24^ Initially, we met regularly as a group to discuss challenges with deciding whether a source should be included or not.^23^ We subsequently met in pairs to complete the resolving of conflicts to improve efficiency. As a significant portion of the conflicts involved studies that focused on the perspectives of sport coaches, administrators, or leadership, we decided to add this to the exclusion criteria. As we began the full text screen, we created a hierarchy of exclusion criteria to limit the number of conflicts in Covidence that appeared because the reviewers chose different reasons for exclusion. Each full text was read by two team members, and if a conflict occurred, the full text was read by at least one additional individual to make a final decision about its inclusion. The first author kept a research diary, minuted every team meeting, and archived email communication to document and describe the decisions we made along the way.^25^

### Data extraction

We collectively developed an extraction data form which captured pertinent information relevant to our research questions.^23^ As with the search strategy and determining inclusion and exclusion criteria, this process was iterative in nature as we discovered additional information we wanted to capture. One team member (OA) read and charted all included articles, while two other team members (JLP and SEC) each read and charted about one half of the references. Any disagreements resulted in a consultation with the wider team to agree on relevant information to be extracted and make amendments to the extraction form as needed. Having one team member read and chart all references added consistency to the review.^25^ Data extracted included the following categories:

- Citation details (authors, date, title)
- Country where research took place
- Study population characteristics (age, race/ethnicity, class) and sample size
- Type and level of sport
- Study aims
- Study design
- Mention of intersectionality
- Gendered environment (pre-sport, training, competition, and/or treatment)
- Gendered environmental features (socio-cultural and material)
- Mention of relationship between sports injury and environment
- Main findings

### Data synthesis

To address the research questions, we descriptively summarized and presented the data in text and table format. To summarize the gendered environmental features in the included papers, the last author (SEC) qualitatively described the data using a thematic survey approach.^45,46^ We did not examine the included studies for quality or undertake a formal thematic analysis.

## RESULTS

The electronic database search identified 17,148 studies for potential inclusion (Figure 2). We removed 6204 duplicates and excluded 10,090 studies after title/abstract screening. Of the 854 references that underwent full text screening, 73 were included in this scoping review (Table 2). Figure 2 shows the total number of studies at each stage of the process, and the reasons for the excluded studies.

**Table 2.** Characteristics of studies included in the scoping review

| **Table 2** Characteristics of studies included in the scoping review |  |  |  |  |  |  |
| --- | --- | --- | --- | --- | --- | --- |
| **Author(s)** | **Year** | **Country** | **Participant age** | **Sport** | **Level of sport** | **Sport environment (pre-sport, training, competition, treatment, or other)** |
| Adjepong<sup>59</sup> | 2017 | USA | Median = 28<br>Range = 18-36 | Rugby sevens | Competitive amateur or club | Competition |
| Ahmad<sup>60</sup> | 2011 | UK | Range = 18-26 | Football | National team/play internationally | Training and competition |
| Aiello-Vaisberg, Kopanakis, & da Silva<sup>61</sup> | 2021 | Brazil | Range = 20-35 | Football | Professional | Training and competition |
| Allison<sup>47</sup> | 2021 | USA | Mean = 31<br>Range = 24-39 | Football | Professional | Training and competition |
| Angelova-Igova & Naydenova<sup>55</sup> | 2023 | Bulgaria | Range = 18-39 | Football | Professional | Training and competition |
| Berg, Duffy, & DuBois<sup>62</sup> | 2022 | USA | Not provided | Rugby sevens | University/college | Competition |
| Biram<sup>63</sup> | 2021 | Brazil | Not provided | Football | Professional | Training and competition |
| Bowes & Kitching<sup>64</sup> | 2021 | UK | Not provided | Golf | Professional | Competition |
| Chahardovali & McLeod<sup>65</sup> | 2023 | USA | Not provided | Softball and football | Professional | Other |
| Comley<sup>66</sup> | 2016 | USA | Range = 18-50 | Surfing | Recreational | Training and competition |
| Cortes de Oliveira, Altmann, & Marques<sup>67</sup> | 2019 | Brazil | ≥ 18 years | Rugby sevens | National team/play internationally | Pre-sport, training, and competition |
| Costa, Kochanek, & Erickson<sup>68</sup> | 2023 | Brazil | Mean = 26 | Skateboarding | Recreational to professional | Training and competition |
| Couch et al.<sup>69</sup> | 2023 | USA | Mean = 48<br>Range = 35-56 | Athletics | Olympic | Pre-sport, training, and competition |
| Cox & Thompson<sup>70</sup> | 2000 | New Zealand | Range = 18-33 | Football | Professional | Pre-sport, training, and competition |
| Culvin<sup>71</sup> | 2021 | England | Range = 20-34 | Football | Professional | Pre-sport, training, and competition |
| Culvin & Bowes<sup>72</sup> | 2021 | England | Range = 20-34 | Football | Professional | Other |
| de Moura et al.<sup>73</sup> | 2023 | Brazil | Not provided | Rugby sevens | National team/play internationally | Training and competition |
| Dogusan & Kocak<sup>74</sup> | 2021 | Turkey | Range = 18-31 | Ice hockey | National team/play internationally | Training and competition |
| Dortants & Knoppers<sup>75</sup> | 2016 | Netherlands | Range = 18-30 | Boxing | Competitive amateur or club | Training and competition |
| Ekengren et al.<sup>76</sup> | 2020 | Sweden | Mean = 31<br>Range = 28-34 | Handball | Professional | Pre-sport, training, and competition |
| Engh, Settler, & Agergaard<sup>77</sup> | 2017 | Denmark | Not provided | Football | Professional | Training and competition |
| Ezzell<sup>78</sup> | 2009 | USA | Not provided | Rugby sevens | University/college | Competition |
| Fisher & Dennehy<sup>79</sup> | 2015 | Brazil | Not provided | Football | Professional | Pre-sport, training, and competition |
| George<sup>80</sup> | 2005 | USA | Not provided | Football | University/college | Training and competition |
| Gill<sup>81</sup> | 2007 | Britain | Not provided | Rugby sevens | Competitive amateur or club | Training and competition |
| Halbert<sup>82</sup> | 1997 | USA | Mean = 28<br>Range = 23-34 | Boxing | Professional | Training and competition |
| Hanis-Martin<sup>83</sup> | 2006 | USA | Range = 21-32 | Basketball | Professional | Training, competition, and treatment |
| Harkness<sup>84</sup> | 2012 | Iraq | Not provided | Football | University/college | Pre-sport and competition |
| Harris<sup>85</sup> | 2001 | England | Not provided | Football | University/college | Training and competition |
| Haselwood et al.<sup>57</sup> | 2005 | USA | Range = 18-23 | Basketball, volleyball, softball, athletics, tennis, football | University/college | Training and competition |
| Horan et al.<sup>86</sup> | 2024 | Ireland | Mean = 27 | Football | Professional | Training and competition |
| Kavoura et al.<sup>87</sup> | 2018 | Finland | Median = 30<br>Range = 20-49 | Judo | National team/play internationally | Training and competition |
| Khetan et al.<sup>58</sup> | 2020 | Nepal | Mean = 26<br>Range = 18-30 | Basketball, badminton, volleyball, cricket, table tennis, football | Recreational to professional | Training and competition |
| Knez, Benn, & Alkhaldi<sup>88</sup> | 2014 | Qatar | Not provided | Football | Professional | Pre-sport, training, and competition |
| Knijnik<sup>89</sup> | 2015 | Brazil | Range = 16-27 | Football | Competitive club to national team | Training and competition |
| Knijnik, Horton, & Cruz<sup>90</sup> | 2010 | Brazil | Range = 18-23 | Surfing | Professional | Pre-sport, training, and competition |
| Kolnes<sup>91</sup> | 1995 | Norway | Late teens to mid-thirties | Handball, football, athletics, cross-country skiing | National team/play internationally | Pre-sport, training, and competition |
| Kristiansen, Broch, & Pedersen<sup>92</sup> | 2014 | USA | Range = 18-37 | Football | Professional | Pre-sport, training, and competition |
| Lafferty & McKay<sup>93</sup> | 2004 | Australia | Not provided | Boxing | Recreational and competitive club | Training |
| Lauer et al.<sup>94</sup> | 2018 | USA | Mean = 21 | Basketball, cross country, softball, volleyball | University/college | Training and competition |
| Litchfield<sup>48</sup> | 2011 | Australia | Early 30s to early 50s | Field hockey | Competitive amateur or club | Training and competition |
| Litchfield & Dionigi<sup>49</sup> | 2013 | Australia | Range = 47-73 | Field hockey | Competitive amateur or club | Competition |
| Mann & Krane<sup>50</sup> | 2018 | USA | Range = 19-23 | Basketball, crew, field hockey, golf, softball, athletics | University/college | Competition |
| Martos-Gracia et al.<sup>51</sup> | 2023 | Spain | Range = 25-30 | Football, futsal | Professional | Pre-sport, training, and competition |
| McGannon et al.<sup>52</sup> | 2019 | Canada | Mean = 28<br>Range = 26-31 | Boxing | National team/play internationally | Training and competition |
| McGannon & McMahon<sup>95</sup> | 2022 | North America | Mean = 41<br>Range = 39-42 | Half-marathon, marathon, ultra-marathon | Competitive amateur or club | Training and competition |
| McGannon, McMahon, & Gonsalves<sup>96</sup> | 2018 | North America | Mean = 41<br>Range = 37-42 | Triathlon, half marathon, full marathon | National team/play internationally | Training and competition |
| McGinnis & Gentry<sup>97</sup> | 2006 | USA | Range = 26-75 | Golf | Recreational | Training and competition |
| McGinnis, Gentry, & McQuillan<sup>98</sup> | 2009 | USA | Range = 26-70 | Golf | Recreational | Training and competition |
| McGinnis, McQuillan, & Chapple<sup>99</sup> | 2005 | USA | Range = 26-70 | Golf | Recreational | Training and competition |
| Mean & Kassing<sup>100</sup> | 2008 | USA | Not provided | Football | Professional | Training and competition |
| Mirehie<sup>101</sup> | 2022 | USA | Range = 27-61 | Marathon, biathlon, triathlon | Recreational | Training and competition |
| Mitchell, Allen-Collinson, & Evans<sup>102</sup> | 2016 | England | Not provided | Golf | Recreational and competitive club | Training |
| Nuhrat<sup>103</sup> | 2022 | Turkey | Not provided | Football | Recreational and competitive club | Pre-sport, training, and competition |
| Oliveira Souza & Capraro<sup>104</sup> | 2020 | Brazil | Not provided | Football | National team/play internationally | Training and competition |
| Organista & Kossakowski<sup>53</sup> | 2024 | Poland | Range = 21-33 | Football | Professional | Training and competition |
| Ozturk & Koca<sup>105</sup> | 2021 | Turkey | Range = 15-23 | Football | Competitive amateur or club | Pre-sport, training, and competition |
| Pelak<sup>106</sup> | 2002 | USA | Not provided | Ice hockey | University/college | Training and competition |
| Rima, Weishaar, & McGladrey<sup>107</sup> | 2019 | USA | >18 years | Volleyball, basketball | University/college | Training and competition |
| Ruggiero & Lattin<sup>108</sup> | 2008 | USA | Mean = 23<br>Range = 18-24 | Basketball | University/college | Training and competition |
| Russell & Lemon<sup>109</sup> | 2012 | Australia | Range = 18-26 | Snowboarding | National team/play internationally | Training |
| Ryan<sup>110</sup> | 2006 | New Zealand | Range = 39-55 | Field hockey | Competitive amateur or club | Competition |
| Sanz-Marcos<sup>111</sup> | 2021 | Spain | Range = 27-41 | Surfing | Recreational | Training |
| Schinke et al.<sup>112</sup> | 2019 | Canada | Mean = 28 | Boxing | National team/play internationally | Training |
| Scraton, Caudwell, & Holland<sup>56</sup> | 2005 | UK | Range = 20-35 | Football | Competitive amateur or club | Pre-sport, training, and competition |
| Scraton et al.<sup>113</sup> | 1999 | England, Germany, Norway, Spain | Not provided | Football | Professional | Pre-sport, training, and competition |
| Sisjord<sup>114</sup> | 2009 | Norway | Range = 18-28 | Snowboarding | Recreational to international/national level | Training and competition |
| Sisjord<sup>115</sup> | 2013 | Norway | Range = 18-31 | Snowboarding | Recreational to international/national level | Training and competition |
| Skogvang & Fasting<sup>116</sup> | 2013 | Norway | Range = 19-35 | Football | Competitive club to national team | Training and competition |
| Sohal, Gervis, & Rhind<sup>117</sup> | 2013 | India | Mean = 29<br>Range = 18-21 | Marathon, tennis, gymnastics, swimming, football, cricket, netball, hockey | National team/play internationally | Training and competition |
| Tamminen, Holt, & Neely<sup>118</sup> | 2013 | Canada | Range = 18-23 | Athletics, swimming, long-distance running, basketball | National team/play internationally | Training and competition |
| Tomlinson & Yorganci<sup>54</sup> | 1997 | Great Britain | Not provided | Athletics | Recreational to international/national level | Training and competition |
| Young<sup>119</sup> | 2004 | Canada | Range = 18-29 | Skateboarding, snowboarding | Recreational to professional | Training |

### Participant and study characteristics

The number of participants across all included studies was approximately 1600; studies that included a description of their sample size reported participant numbers ranging from 4 to 300 individuals. More than half of the studies (n = 52) were conducted in North America and Europe. Twenty-nine studies described aspects of the cultural or ethno-racial demographics of their sample. Seven studies described the sexual identities of their sample.^47–53^ The most commonly studied sport was football (soccer). The level of play of the athletes in the included studies varied. Professional athletes were most often studied; recreational and club level athletes were least often the target population. More than half of the included studies discussed the intersection of gender with other identities (e.g., race, sexuality) in their results and/or discussion sections beyond simply including those identities as characteristics of the study sample. Most studies used qualitative methods, with only 5 of the 73 studies (7%) employing quantitative methods.^54–58^ Interviews were the most common qualitative method, but observation, focus groups, ethnography and arts-based methods were also reported. See Table 2 for a detailed description of the included studies.

### Gendered sport environments

When considering the four sport environments we identified in our gendered environmental approach,^22^ most studies focused on the Training and Competition environments, very few touched on the Pre-sport environment, and only one included information relevant to the Treatment environment. We identified two studies that described environmental features such as labour policies that did not neatly fit into any of our four proposed sport environments; we categorized these as “other”.^65,72^

Only 19 of the 73 included studies specifically mentioned a relationship between gendered sport environments and injury.^54,55,59,62,63,71,72,83,85,86,93,100,101,109,114,115,117–119^ Most of these 19 studies contained minimal discussion on the topic, and any mention of injury was restricted to one or two direct quotes from an athlete. Most commonly, the lack of adequate resources (first aid equipment, poor field conditions, lack of health insurance coverage) in relation to injury risk and experiences were described.

We identified three repeating patterns within the included studies when summarizing the gendered sport environments that women athletes encounter.

#### 1) “Ladies tees” and “tarts”: Embedded stereotypes that devalue women and women’s sport

Fifty-five studies reported gender stereotypes embedded within women’s sport environments that worked to normalize women athletes as occupying a position secondary to men. Studies described how negative stereotypes that cast women as fragile, ‘dainty,’ unfit for sport, or unserious about sport permeated everyday sport experiences,^54,67,73,84,85,87–89,91,92,97–100,109,111,112^ including ingrained ideas about women as biologically inferior to men.^47,52,54,59,63,64,86,87,93,100,102,103,119^ Such stereotypical norms effectively peripheralize women within the sport setting, while benchmarking men as ‘true’ athletes.^100^

Institutionalized practices normalized women’s secondary position, such as prioritizing men’s opportunities even when women’s teams achieved better results^55,73,85,105,115^ or not recognizing women’s achievements.^105,112^ These disenfranchising stereotypes were entrenched in everyday language across sports, such as learning to surf by body-boarding (lying prone on a board) being called ‘women’s surfing’^90^ or ‘futebol femenino’ (women’s football) being used as a ‘lesser than’ term.^89^ Outright sexism, discrimination, and harassment were commonly reported,^48,52,54,58,66,68,72,82,83,89,91,93,97,101–103,106,108,119^ such as women runners being catcalled.^101^ Two studies described gender stereotypes specifically related to coaches, where athletes believed a “protective father-figure would be best”^107^ and male coaches were more direct in their communication style.^57^ Other studies demonstrated how characteristics such as race and age intersect with these stereotypes in the sport environment to doubly disadvantage minoritized sportswomen, from exposure to everyday microaggressions to overt verbal abuse and discrimination.^56,60,69,108,110^

Studies additionally reported how gender stereotypes (e.g., nurturing qualities) further condition women’s participation on providing additional gendered labour, such as volunteer work or fan engagement whereas their male counterparts are not expected to do the same.^47,63,92^ Such “unwaged inspirational work” was tied to pressure on women to serve as role models,^65^ and relates to another norm that women athletes should be grateful for their opportunities despite the myriad disadvantages and barriers they face compared with men.^47,65,72,105^ The sport environment taxes women athletes with expending affective labour, where their time could be better spent training or recovering.^47,65^

#### 2) “Play like a man and perform like a lady off the pitch:” Restricting or redefining femininities?

Forty-five studies described ways that the sport environment both reproduces restrictive behavioural and bodily gender norms for women, while in other ways acts as a counter-hegemonic space for more flexible femininities. One of the features of this was compulsory heterosexuality, and in particular ensuring the *appearance* of heterosexuality.^53,59,70,73,78,80–82,92,93,98,113,116^ This dimension of the gendered sport environment can impose another form of invisible labour where women feel compelled to manage how their gender and sexuality are ‘read’, both on and off the field of play.^59,71^ Such norms could hold women back. For example, in golf, McGinnis et al. (2009) found that being ‘too good’ was seen to compromise femininity, and so some women were hesitant to be too competitive. Other studies described how the sport environment created what could be described as ‘contingent’ femininities, meaning women athletes could ‘acceptably’ exhibit traditionally masculine traits if counter-balanced with traditional feminine qualities.^62,91,92,103,105^ Studies also reported instances where sport environments rewarded the adoption of conventionally masculine traits by women, including examples from boxing,^75^ handball,^76^ rugby,^78^ and surfing.^111^ Still, such traits could be contentious off-the-field with Gill (2007), for example, reporting disapproval of women rugby players being loud, boisterous, and conspicuously consuming alcohol. There were racialized dimensions to ‘acceptable’ sport femininities, with studies reporting how Black women athletes could be expected to embody stereotypes about ‘natural’ strength^77^ or speed.^56^ Still, in some cases, sport environments created space for flexible femininities and were more open to women participating on their own gendered terms.^50–52,101,119^ In an example from the Canadian women’s boxing team, McGannon et al. (2019) reported how valuing aggression and skill destabilized normative femininity.

Other studies reported how sport environments regulate women’s bodily presentation through clothing and physique norms. The literature described how physique operated as a representation of a ‘real’ athlete,^75,79^ and for women this often entailed maintaining a heterosexualised feminine aesthetic.^52,80,90^ Some women exaggerated their physical feminine presentation to avoid any ‘gender trouble’^89^ or engaged in body manipulation – through dieting, or excessive exercise – to conform to an idealized female athlete body.^89,94,103,105^ Clothing was a way to manage feminine bodily representations. Several studies reported how clothing was both a way to demonstrate traditional femininity (e.g., “wear skirts in competition so you could ‘tell they were women”)^75,82^ as well as to ‘cover’ femininity to avoid sexualised attention.^68,90,105^ Negotiating different feminine presentations and styles could be a means of acceptance (or exclusion) in sport.^114^ In boxing, Lafferty and McKay (2004) reported how women who wore revealing clothes were not taken seriously and sexualized by male boxers, while at the same time men were free to take their shirt off to box without social sanction. Clothing and uniforms could also function as exclusionary devices by being incompatible with certain cultural or religious codes for some women.^60,88^

While the majority of gendered environments described in the included studies resulted in women facing barriers or challenges and being disadvantaged, there were exceptions where positive outcomes were noted. A gendered environment can result in an inclusive and supportive team setting where athletes feel accepted for who they are and do not need to expend energy to present themselves in a ‘socially acceptable’ way.^50,51^

#### 3) “Training on the men’s pitch was seen as a ‘treat:’” Gendered inequities in opportunities and resources

Forty-six studies reported ways that gendered inequities were structurally embedded in the material conditions of women’s sport environments. Core among these were systemically poor labour conditions, including inadequate remuneration and gendered wage inequality,^47,55,61,73,76,92,104^ which can result in athletes having to work secondary jobs outside of sport.^55,86,104^ Horan et al. (2024) describe how this dual employment can impede athlete technical and tactical development by taking time and energy away from sport. This can be compounded by the additional burden of some athletes having to manage their own training and travel.^117^ Women also described being eligible for less prize money than men,^68,90^ and in some cases personally covering some of their own expenses.^55,117^ Studies also reported precarious labour conditions and contracts,^47^ including termination if pregnant^55^ or inadequate maternity protection,^72,76^ and insufficient benefits packages.^47,63^ Related to the devaluing of women’s time in Pattern 1, women athletes could also be subject to ‘extracurricular’ labour, additional role expectations, and obligations that were neither outlined in their contracts nor paid.^65^

Studies across a range of different sports reported subpar training facilities, with women relegated to second-rate facilities or having limited access to facilities.^47,56,63,71,74,79,81,82,85,86,93,103–105^ Culvin (2021) reported in football how ‘training on the men’s pitch was regarded as a “treat“’, while Biram et al. (2021) observed how women’s relegation to a secondary pitch afforded a surface upon which technical training was not possible.^63^ These inequities extended to inadequate access to specialist support staff, such as sport psychologists, high-quality medical care, and strength and conditioning professionals.^69,86,117^ There were even examples of only the men’s teams being allocated first aid kits^85^ or getting preferential accommodation over the women’s teams.^117^ Inequities were further embossed in the mundane aspects of sport participation, including inadequate toilet provision^99,105^ and small dressing rooms or no dressing rooms at all.^104,105^

## DISCUSSION

We set out to identify the extent of the current literature that describes the gendered aspects of women’s organized sports environments which are higher risk for ACL injury. To our knowledge, this is the first time describing the breadth of the literature available in this area, and the first attempt to focus on potential links between injuries and the gendered socio-cultural and material aspects of women’s sport environments. Our results uncovered 73 studies, with only about a quarter of those containing any explicit mention of injury in relation to the gendered sport environment. In-depth discussion of injury risk, experiences, and outcomes emanating from women’s gendered sport environments was largely absent and not the primary focus of any of the included studies. The gendered environments in the included studies mapped well onto our existing model,^22^ with the exception of two studies which looked at issues that transcend a single environment (e.g., labour contracts).^65,72^

While very few studies mentioned injury in relation to the gendered environments found in women’s sport, the extensive description of those environments and how they act as barriers and disadvantage women appears to support our contention that sports injury should be considered a bio-*social* phenomenon, with both biological and social influences. Below, we outline some possible theoretical connections between the women’s sport environments described in the included articles and their injury risk and experiences as examples. First, women being thought of as fragile and dainty could plausibly impact injury risk if this results in coaches, parents, or athletes themselves thinking they need to hold back during training. This could mean missing out on the opportunity to train at the appropriate intensities needed to adequately prepare them for the physical demands of their sport. Second, many of the gendered aspects of women’s sport environments described in this scoping review could plausibly lead to decreased time and physical and cognitive energy availability for adequate training and recovery. This could lead to increased injury risk and/or delay in recovery from an injury. For examples, we highlight the additional gendered labour often expected of women athletes but not men; the invisible labour of women needing to keep up ‘acceptable’ feminine appearances by dressing and behaving in certain ways; and the need for women to take on secondary paying jobs because of inadequate sport income. Third, the existence of outright discrimination and harassment could lead to women avoiding certain locations or not training at certain times of day,^120^ potentially missing out on critical training opportunities to improve fitness and physical skills and decrease injury risk. Perhaps the most obvious link to injury is the gendered inequities in opportunities and resources such as provision of less-than-ideal training facilities, inadequate professional and medical support, and inequitable travel conditions and accommodations. Poor field conditions (e.g., uneven surfaces) may contribute to injury,^121^ and inadequate care immediately following an injury or in rehabilitation has the potential to delay return to sport and affect future player availability.^122^ Long travel times and poor accommodations result in physical and mental fatigue for players which can translate into increased injury risk during play.^123^

Some organizations are recognizing these gendered inequities and are taking action to improve the conditions in women’s sport. For example, FIFPRO recently released a comprehensive guide for players and staff to manage athlete pregnancy and return to sport;^124^ the Danish men’s football team refused a pay raise to ensure the women’s team was paid equitably;^125^ and we recently developed an evidence-based online exhibition that is being used in high-performance sport to increase awareness of gendered environments and empower sport staff to take action.^126^ This scoping review confirmed the existence of gendered aspects of women’s sport across diverse settings from football to surfing to field hockey. It is clear that gendered conditions exist for women which can get in the way of them maximizing their potential; the imperative next steps are to go beyond describing the environment to more deeply exploring the relationship with injury, and subsequently taking required action to improve sport conditions for women.

### Limitations, strengths, and future research

As we needed to balance the necessity to conduct a thorough search with feasibility and the resources of our team,^23–25^ we did not review the reference lists of relevant literature we found or conduct a focused grey literature search. We did not critically appraise the final articles for quality or risk of bias, as our primary purpose was to gain general knowledge of the scope of the literature in this topic area. Therefore, our reporting of how the 73 studies fall into the three patterns we identified should be considered with that in mind.^127^ On the other hand, the multidisciplinary social science and clinical nature of our research team adds rigour to our study by providing varied expertise that ensured comprehensive mapping and reporting of the findings.^128^

We excluded many studies that focused on the gendered environments experienced by coaches, administrators, and medical staff. There would be value in summarizing the evidence outlining their experiences, and how it could relate to the injury risk and experiences of athletes. We also focused on adult women, but since many injuries, including ACL rupture, occur at a younger age there is a need to scrutinize the gendered environments encountered by youth athletes.

## CONCLUSION

This scoping review provides a snapshot of the currently available literature describing gendered aspects of women’s sport environments that are higher risk for ACL injury. Existing literature describes a range of gendered inequities that exist for women in their sport environments; however, there has been no concerted effort to date to link those gendered environmental factors to ACL injury. Such research is needed if we are serious about eliminating the ACL injury rate disparity between women and men.

## Data Availability

All data produced in the present work are contained in the manuscript

## Acknowledgements

Thank you to Sheree Bekker for her contributions to the project.

## Appendices

## Appendix A Included Sports

Appendix A: Included Sports
| <u>Summer</u> | <u>Winter</u> |
| --- | --- |
| Athletics/track and field | Biathlon |
| Badminton | Ice hockey |
| Basketball | Skating |
| Boxing | Skiing |
| Breaking (Breakdancing) | Ski jumping |
| Fencing | Ski mountaineering |
| Football | Snowboarding |
| Golf |  |
| Gymnastics |  |
| Handball |  |
| Hockey |  |
| Judo |  |
| Modern pentathlon |  |
| Rugby sevens |  |
| Skateboarding |  |
| Surfing |  |
| Table tennis |  |
| Taekwondo |  |
| Tennis |  |
| Volleyball (beach and indoor) |  |
| Weightlifting |  |
| Wrestling |  |

## Appendix B Search strategy

1 women/
2 female/
3 wom?n*.tw,kf.
4 female*.tw,kf.
5 girl*.tw,kf.
6 femme*.tw,kf.
7 feminin*.tw,kf.
8 or/1-7 [female search]
9 exp sports/
10 exp organizations/
11 9 and 10
12 ((organiz* or federation or committee or (governing adj1 body) or professional or recreation* or competitive or club* or team*) and (sport* or athletic*)).tw,kf.
13 ((organiz* or federation or committee or (governing adj1 body) or professional or recreation* or competitive or club* or team*) and basketball).tw,kf.
14 ((organiz* or federation or committee or (governing adj1 body) or professional or recreation* or competitive or club* or team*) and (football or soccer)).tw,kf.
15 ((organiz* or federation or committee or (governing adj1 body) or professional or recreation* or competitive or club* or team*) and gymnastics).tw,kf.
16 ((organiz* or federation or committee or (governing adj1 body) or professional or recreation* or competitive or club* or team*) and hockey).tw,kf.
17 ((organiz* or federation or committee or (governing adj1 body) or professional or recreation* or competitive or club* or team*) and (handball or lacrosse or rugby)).tw,kf.
18 ((organiz* or federation or committee or (governing adj1 body) or professional or recreation* or competitive or club* or team*) and wrestling).tw,kf.
19 ((organiz* or federation or committee or (governing adj1 body) or professional or recreation* or competitive or club* or team*) and (ski or skiing)).tw,kf.
20 or/11-18 [sport organizations]
21 gender identity/ or gender role/
22 (gender* or sex*).tw,kf.
23 or/21-22 [gender]
24 8 and 20 and 23 [female, gender, sports]
25 limit 24 to english language

